# Antiviral drugs in hospitalized patients with COVID-19 - the DisCoVeRy trial

**DOI:** 10.1101/2021.01.08.20248149

**Authors:** Florence Ader, Nathan Peiffer-Smadja, Julien Poissy, Maude Bouscambert-Duchamp, Drifa Belhadi, Alpha Diallo, Christelle Delmas, Juliette Saillard, Aline Dechanet, Noémie Mercier, Axelle Dupont, Toni Alfaiate, François-Xavier Lescure, François Raffi, François Goehringer, Antoine Kimmoun, Stéphane Jaureguiberry, Jean Reignier, Saad Nseir, François Danion, Raphael Clere-Jehl, Kévin Bouiller, Jean-Christophe Navellou, Violaine Tolsma, André Cabie, Clément Dubost, Johan Courjon, Sylvie Leroy, Joy Mootien, Rostane Gaci, Bruno Mourvillier, Emmanuel Faure, Valérie Pourcher, Sébastien Gallien, Odile Launay, Karine Lacombe, Jean-Philippe Lanoix, Alain Makinson, Guillaume Martin-Blondel, Lila Bouadma, Elisabeth Botelho-Nevers, Amandine Gagneux-Brunon, Olivier Epaulard, Lionel Piroth, Florent Wallet, Jean-Christophe Richard, Jean Reuter, Thérèse Staub, Maya Hites, Marion Noret, Claire Andrejak, Gilles Peytavin, Bruno Lina, Dominique Costagliola, Yazdan Yazdanpanah, Charles Burdet, France Mentre, on behalf of the DisCoVeRy study group

## Abstract

**Background:** Lopinavir/ritonavir, lopinavir/ritonavir-interferon (IFN)-β-1a and hydroxychloroquine efficacy for COVID-19 have been evaluated, but detailed evaluation is lacking.

**Objective:** To determine the efficacy of lopinavir/ritonavir, lopinavir/ritonavir-IFN-β-1a, hydroxychloroquine or remdesivir for improving the clinical, virological outcomes in COVID-19 inpatients.

**Design:** Open-label, randomized, adaptive, controlled trial.

**Setting:** Multi-center trial with patients from France.

**Participants:** 583 COVID-19 inpatients requiring oxygen and/or ventilatory support

**Intervention:** Standard of care (SoC, control), SoC plus lopinavir/ritonavir (400 mg lopinavir and 100 mg ritonavir every 12h for 14 days), SoC plus lopinavir/ritonavir plus IFN-ß-1a (44 μg of subcutaneous IFN-ß-1a on days 1, 3, and 6), SoC plus hydroxychloroquine (400 mg twice on day 1 then 400 mg once daily for 9 days) or SoC plus remdesivir (200 mg intravenously on day 1 then 100 mg once-daily for hospitalization duration or 10 days).

**Measurements:** The primary outcome was the clinical status at day 15, measured by the WHO 7-point ordinal scale. Secondary outcomes included SARS-CoV-2 quantification in respiratory specimens and safety analyses.

**Results:** Adjusted Odds Ratio (aOR) for the WHO 7-point ordinal scale were not in favor of investigational treatments: lopinavir/ritonavir *versus* control, aOR 0.83, 95%CI, 0.55 to 1.26, P=0.39; lopinavir/ritonavir-IFN-β-1a *versus* control, aOR 0.69, 95%CI, 0.45 to 1.04, P=0.08; hydroxychloroquine *versus* control, aOR 0.93, 95%CI, 0.62 to 1.41, P=0.75. No significant effect on SARS-CoV-2 RNA clearance in respiratory tract was evidenced. Lopinavir/ritonavir-containing treatments were significantly associated with more SAE.

**Limitations:** Not a placebo-controlled, no anti-inflammatory agents tested.

**Conclusion:** No improvement of the clinical status at day 15 nor SARS-CoV-2 RNA clearance in respiratory tract specimens by studied drugs. This comforts the recent Solidarity findings.

**Registration:** NCT04315948.

**Funding:** PHRC 2020, Dim OneHealth, REACTing

## Introduction

The new severe acute respiratory syndrome coronavirus 2 (SARS-CoV-2)-associated coronavirus disease 2019 (COVID-19) emerged in China in December 2019, subsequently causing a worldwide pandemic(1, 2).

While waiting for specific therapeutic treatments and/or vaccines against SARS-CoV-2, efforts initially focused on repurposing drugs that have previously shown broad-spectrum antiviral activity against other coronaviruses. Lopinavir/ritonavir, type I interferon (IFN), hydroxychloroquine, and remdesivir were among the first investigational treatments to be tested on the basis of their *in vitro* activity against SARS-CoV-2(3-9). Activity of combined anti-HIV agent lopinavir/ritonavir against novel coronaviruses occurs *via* inhibition of 3-chymotrypsin-like protease-dependent proteolysis preventing cleavage into viral proteins(3, 4). Activity of hydroxychloroquine occurs *via* affecting early stage of viral replication through increased endosomal pH resulting in inhibition of virus-endosome fusion(7-9). Activity of type I IFN is mediated by the heightening of antiviral cellular immune defenses(10). In addition, it was hypothesized that hydroxychloroquine and type I IFN may have intrinsic immunomodulatory effects(10-12). Remdesivir is an adenosine analogue, which incorporates into nascent viral RNA chains and results in pre-mature termination(13).

The DisCoVeRy trial is a phase 3 multicenter, open-label, randomized, adaptive, controlled trial to evaluate the clinical and the virological efficacy, as well as the safety, of lopinavir/ritonavir, lopinavir/ritonavir plus IFN-β-1a, hydroxychloroquine, and remdesivir as compared with standard of care in adults hospitalized for COVID-19 (14). The DisCoVeRy trial is part of the international Solidarity trial consortium sponsored by the World Health Organization (WHO). Interim results of the Solidarity trial were recently published. They were restricted to the analysis of in-hospital mortality, need for mechanical ventilation and time to hospital discharge. No treatment exhibited a significant effect on these critical outcomes when compared to standard of care(15). The DisCoVeRy trial provides more detailed data on safety and clinical and laboratory surrogates, in particular virological data. We report here the results from the lopinavir/ritonavir, lopinavir/ritonavir plus IFN-β-1a and hydroxychloroquine arms, which were prematurely stopped. The remdesivir arm is continuing and currently enrolling.

## Methods

### Trial design and oversight

DisCoVeRy is an open-label, adaptive, multicenter, randomized controlled trial which evaluates the efficacy and safety of drugs in adults hospitalized for COVID-19.

Sponsored by the Institut national de la santé et de la recherche médicale (Inserm, France), the trial was approved by the Ethics Committee (CPP Ile-de-France-III, approval #20.03.06.51744). Written informed consent was obtained from all included participants or their legal representative if they were not able to consent. The trial was conducted in accordance with the Declaration of Helsinki and national laws and regulations and declared on the clinicatrials.gov registry (NCT 04315948) and on the European Clinical Trials Database (2020-000936-23). The present analysis is based on the protocol v7.0 of April, 5^th^ 2020,(14) with two secondary outcomes added in protocol v9.0 of June, 29^th^ 2020.

### Participants

Eligible participants were adults (≥ 18-year old) hospitalized with a PCR-proven (< 72 hours) SARS-CoV-2 infection and pulmonary rales or crackles with a peripheral oxygen saturation ≤ 94%, or requirement of supplemental oxygen. Exclusion criteria included a contraindication to any of the investigational treatment or their use within 29 days before randomization; elevated liver enzyme > 5 times the upper limit, an estimated glomerular filtration rate below 30 mL/min, pregnancy or breast-feeding. Full inclusion and exclusion criteria are presented in the Supplementary Appendix.

### Interventions and randomization

Participants were randomly assigned in a 1:1:1:1:1 ratio through computer-generated blocks of various sizes and stratified by administrative region and severity of illness at enrolment (moderate disease: hospitalized participants receiving low-flow supplemental oxygen or not requiring oxygen; severe disease: hospitalized participants requiring non-invasive ventilation or high-flow oxygen devices, invasive mechanical ventilation or ECMO). Randomization was implemented in the electronic Case Report Form to ensure appropriate allocation concealment. Investigational arms were standard of care (SoC, control), SoC plus lopinavir/ritonavir (tablets or oral suspension in nasogastric tube of 400 mg lopinavir and 100 mg ritonavir administered every 12h for 14 days), SoC plus lopinavir/ritonavir plus IFN-ß-1a (44 μg of subcutaneous IFN-ß-1a on days 1, 3, and 6), SoC plus hydroxychloroquine (200 mg tablets adjusted to 400 mg twice on day 1 as a loading dose followed by 400 mg once daily for 9 days)(16) or SoC plus remdesivir (200 mg intravenously on day 1 as a loading dose followed by 100 mg once-daily for hospitalization duration or 10 days). Other supportive treatments, such as glucocorticoids or immunomodulatory agents, were allowed in all arms.

### Clinical and laboratory monitoring

Participants were assessed daily while hospitalized, and at days 3, 5, 8, 11, 15±2 and 29±3 if they were withdrawn at these dates. Clinical data, concomitant medications, adverse events (AEs) and measurements for blood cell counts, serum creatinine and liver aminotransferases were collected. Nasopharyngeal swab (NPS) and lower respiratory tract (LRT, *i*.*e*. bronchoalveolar lavage, protected specimen brushing or bronchial aspiration) specimens were collected for SARS-CoV-2 real-time (RT) PCR.

### Outcomes measures

The primary outcome measure was the clinical status at day 15 as measured on the 7-point ordinal scale of the WHO Master Protocol (v3.0, March 3, 2020): 1. Not hospitalized, no limitation on activities; 2. Not hospitalized, limitation on activities; 3. Hospitalized, not requiring supplemental oxygen; 4. Hospitalized, requiring supplemental oxygen; 5. Hospitalized, on non-invasive ventilation or high flow oxygen devices; 6. Hospitalized, on invasive mechanical ventilation or ECMO; 7. Death.

Secondary efficacy outcome measures included the clinical status at day 29 and the time to an improvement of 2 categories as measured on the 7-point ordinal scale or hospital discharge until day 29, the time to National Early Warning Score 2 (NEWS2) ≤2 or hospital discharge until day 29, the time to hospital discharge until day 29, oxygenation- and ventilator-free days until day 29, 29-day mortality. Normalized SARS-CoV-2 viral loads were qualitatively and quantitatively assessed in respiratory tract specimens (see Supplementary Appendix). Secondary safety outcomes included the cumulative incidence of any grade 3 or 4 AE (coded using the medical dictionary for regulatory affairs, v23.0), or of any serious adverse event (SAE, according to the Division of AIDS (DAIDS, Table for Grading the Severity of Adult and Paediatric Adverse Events, v2.1, July 2017) and the proportion of patients with a premature suspension or discontinuation for any reason of the investigational treatments.

### Sample size calculation

Hypotheses for sample size calculation are presented in Supplementary Appendix.

### Interim analyses

An independent data safety and monitoring board (DSMB) externally reviewed the trial data. For efficacy and futility, the statistical analysis was performed on the primary outcome measure, and was based on the Haybittle-Peto rule(17, 18). Consistently with its role of add-on partner trial, DisCoVeRy periodically transferred data regarding three of its secondary outcomes to the WHO Solidarity trial: in-hospital mortality, time to hospital discharge, time to mechanical ventilation.

On May 25^th^ 2020, following a safety warning on hydroxychloroquine use(19), enrollment in the hydroxychloroquine arm was suspended at the request of the French Agency of drug Security (Agence Nationale de Sécurité du Médicament). On June 13^th^, based on the interim analysis of the Solidarity data, the Solidarity and DisCoVeRy trial DSMBs recommended to definitely stop the hydroxychloroquine arm due to futility. This decision was endorsed by the DisCoVeRy steering committee on June 17^th^. The Solidarity DSMB advised to stop the lopinavir/ritonavir arm due to futility on June, 23^th^. Thereafter, the DisCoVeRy DSMB further advised to stop both the lopinavir/ritonavir-containing arms due to additional safety concern on June, 25^th^. This decision was endorsed by the DisCoVeRy steering committee on June 27^th^ with subsequent interruption on June, 29^th^.

### Statistical analyses

Statistical analyses compared each of the three stopped investigational treatment arms to the control arm. Due to the slowdown of inclusions in the trial between May and July, a single patient was included in the control arm between the hydroxychloroquine arm suspension and the permanent discontinuation of the lopinavir/ritonavir-containing arms. For simplification purpose, only one control group was constituted, which included participants from the control arm included before June 29^th^. The intention-to-treat population included all randomized participants for whom a valid consent form was obtained. The modified intention-to-treat population included participants from the intention-to-treat population who received at least one dose of the treatment allocated by randomization.

Efficacy analyses were performed on the intention-to-treat population; handling of missing data is described in Supplementary Appendix. Safety analyses were performed on modified intention-to-treat population. Analyses were stratified by baseline severity but not by region of inclusion due to a low number of inclusions in some regions; all tests were two-sided with a type-I error of 0.05.

For the 7-point ordinal scale, data were analyzed using a proportional odds model. Time-to-event data were analyzed using a Cox proportional hazard model. An analysis of covariance was performed for the comparison of oxygenation- and ventilator-free days between groups; 29-day mortality and the number of participants with detectable SARS-CoV-2 in the respiratory tract specimens at each time point were analyzed using a Cochran-Mantel-Haenszel test. Log_10_ normalized SARS-CoV-2 load kinetics was analyzed using a mixed-effects linear model with test of treatment effect on slopes. For safety endpoints, non-prespecified statistical comparisons of the proportions of patients with any i) adverse event, ii) grade 3 or 4 adverse event, or iii) serious adverse event between each investigational treatment arm versus control were performed using the Fisher exact test.

### Role of the funding source

The trial was funded by grants from Programme Hospitalier de Recherche Clinique (PHRC-20-0351) (Ministry of Health), from the DIM One Health Île-de-France (R20117HD) and from REACTing, a French multi-disciplinary collaborative network working on emerging infectious diseases. The funding sources had no role in the analysis of the data nor in the decision of publication.

## Results

### Patient’s characteristics at baseline

Between March, 22^nd^ and June, 29^th^, 603 participants were randomized to one of the 4 arms across 32 sites in France and Luxembourg and 583 were evaluable for analysis (Supplementary Figure S1): control arm, n=148; lopinavir/ritonavir arm, n=145; lopinavir/ritonavir plus IFN-ß-1a arm, n=145; hydroxychloroquine arm, n=145. Participants’ baseline characteristics are presented in the Table 1. The median age was 63 years (IQR, 54; 71) and participants were mostly male (n=418, 71.7%). The median time from symptom onset to randomization was 9 days (IQR, 7; 12). The most frequent underlying conditions were obesity (n=166, 28.7%), chronic cardiac disease (n=151, 26.0%) and diabetes mellitus (n=128, 22.0%). At baseline, 211 (36.2%) participants presented a severe disease. Concomitant treatments are presented in Supplementary Table 1. No notable imbalance between groups was observed regarding concomitant anti-inflammatory drug used.

**Table 1.** Baseline characteristics of patients included in the intention to treat population of the present analysis of DisCoVeRy trial. NPS, Nasopharyngeal swabs; LRT, Lower respiratory tract. * denotes variables with missing data. Data on chronic cardiac disease, chronic pulmonary disease, mild liver disease, chronic neurological disorder, active cancer and diabetes mellitus were missing for 2 patients; data on chronic kidney disease were missing for 3 patients; data on auto-inflammatory disease were missing for 1 patient; data on obesity were missing for 5 patients; data on smoking status were missing for 30 patients; data on the time from symptoms onset to randomization were missing for 8 patients; data on BMI were missing for 83 patients; data on randomization site were missing for 1 patient; data on viral load from NPS were missing for 234 patients; data on viral load from LRT specimens were missing for 527 patients; data for lymphocyte count were missing for 90 patients; data for neutrophil count were missing for 136 patients; data on creatinine were missing for 15 patients; data on AST/SGOT were missing for 56 patients; data on ALT/SGPT were missing for 51 patients; data on CRP were missing for 137 patients; data on D-Dimers were missing for 299 patients; data on PCT were missing for 356 patients; data on ferritin were missing for 421 patients. ** moderate disease: hospitalized participants receiving low-flow supplemental oxygen or not requiring oxygen; severe disease: hospitalized participants requiring non-invasive ventilation or high-flow oxygen devices, invasive mechanical ventilation or ECMO

|  | **Overall**<br>**(N=583)** | **Control**<br>**(N=148)** | **Lopinavir/ritonavir (L/r)**<br>**(N=145)** | **Lopinavir/Ritonavir +**<br>**Interferon β-1a (L/r + IFN)**<br>**(N=145)** | **Hydroxychloroquine**<br>**(HCQ)**<br>**(N=145)** |
| --- | --- | --- | --- | --- | --- |
| **Median age — yr [IQR]** | 63 [54-71] | 62 [52-71] | 63 [55-71] | 64 [53-71] | 65 [55-71] |
| **Male sex — no. (%)** | 418 (71.7%) | 105 (70.9%) | 106 (73.1%) | 103 (71.0%) | 104 (71.7%) |
| **Coexisting condition\* — no. (%)** |  |  |  |  |  |
| - Chronic cardiac disease | 151 (26.0%) | 39 (26.4%) | 35 (24.1%) | 36 (25.2%) | 41 (28.3%) |
| - Chronic pulmonary disease | 88 (15.1%) | 31 (20.9%) | 19 (13.1%) | 19 (13.3%) | 19 (13.1%) |
| - Chronic kidney disease (stage 1 to 3) | 24 (4.1%) | 7 (4.7%) | 2 (1.4%) | 5 (3.5%) | 10 (6.9%) |
| - Mild liver disease | 13 (2.2%) | 6 (4.1%) | 3 (2.1%) | 0 (0.0%) | 4 (2.8%) |
| - Chronic neurological disorder<br>(including dementia) | 23 (4.0%) | 6 (4.1%) | 5 (3.4%) | 4 (2.8%) | 8 (5.5%) |
| - Active cancer | 35 (6.0%) | 10 (6.8%) | 8 (5.5%) | 6 (4.1%) | 11 (7.6%) |
| - Auto-inflammatory disease | 26 (4.5%) | 8 (5.4%) | 4 (2.8%) | 9 (6.3%) | 5 (3.4%) |
| - Obesity | 166 (28.7%) | 46 (31.3%) | 36 (24.8%) | 41 (28.7%) | 43 (30.1%) |
| - Diabetes mellitus | 128 (22.0%) | 35 (23.6%) | 35 (24.1%) | 27 (18.9%) | 31 (21.4%) |
| - Current smoker | 18 (3.3%) | 5 (3.5%) | 4 (2.9%) | 5 (3.6%) | 4 (3.0%) |
| **Median time from symptom onset**<br>**to randomization\* — days [IQR]** | 9.0 [7.0-12.0] | 10.0 [7.0-12.0] | 10.0 [7.0-13.0] | 10.0 [7.0-12.0] | 8.0 [7.0-11.0] |
| **Baseline severity of COVID-19\*\* — no. (%)** |  |  |  |  |  |
| - Moderate | 372 (63.8%) | 94 (63.5%) | 94 (64.8%) | 91 (62.8%) | 93 (64.1%) |
| - Severe | 211 (36.2%) | 54 (36.5%) | 51 (35.2%) | 54 (37.2%) | 52 (35.9%) |
| **Randomization site\* — no. (%)** |  |  |  |  |  |
| - ICU | 254 (43.6%) | 64 (43.2%) | 65 (44.8%) | 65 (45.1%) | 60 (41.4%) |
| - Conventional unit (e.g. : infectious<br>disease unit, internal medicine,<br>pneumology) | 328 (56.4%) | 84 (56.8%) | 80 (55.2%) | 79 (54.9%) | 85 (58.6%) |
| **7-point ordinal scale at baseline — no. (%)** |  |  |  |  |  |
| - 3. Hospitalized, not requiring<br>supplemental oxygen | 27 (4.6%) | 8 (5.4%) | 4 (2.8%) | 9 (6.2%) | 6 (4.1%) |
| - 4. Hospitalized, requiring<br>supplemental oxygen | 341 (58.5%) | 84 (56.8%) | 88 (60.7%) | 84 (57.9%) | 85 (58.6%) |
| - 5. Hospitalized, on non-invasive<br>ventilation or high flow oxygen<br>devices | 63 (10.8%) | 21 (14.2%) | 15 (10.3%) | 13 (9.0%) | 14 (9.7%) |
| - 6. Hospitalized, on invasive<br>mechanical ventilation or ECMO | 152 (26.1%) | 35 (23.6%) | 38 (26.2%) | 39 (26.9%) | 40 (27.6%) |
| **Median NEWS-2 at baseline\*,** | 9.0 [7.0-12.0] | 9.0 [7.0-12.0] | 9.0 [7.0-11.0] | 10.0 [7.0-12.0] | 9.0 [6.0-11.0] |
| **median [IQR]** |  |  |  |  |  |
| **Median viral load at baseline, median [IQR]** |  |  |  |  |  |
| - on NPS (log10 copies/10 000 cells) | 2.4 [0.7-3.7]<br>(n=349) | 2.5 [1.1-3.9]<br>(n=87) | 2.4 [0.7-3.6]<br>(n=88) | 2.5 [0.7-3.8]<br>(n=79) | 2.0 [0.7-3.4]<br>(n=95) |
| - on LRT specimens (log10 copies/10 000 cells) | 4.1 [2.8-4.9]<br>(n=56) | 3.6 [2.4-4.5]<br>(n=14) | 4.4 [3.2-4.8]<br>(n=14) | 3.5 [1.0-4.8]<br>(n=10) | 4.3 [3.3-5.5]<br>(n=18) |
| **Biological data at baseline\*, median [IQR]** |  |  |  |  |  |
| - Minimal lymphocytes count (G/L) | 0.9 [0.6-1.2] | 0.9 [0.6-1.4] | 0.8 [0.6-1.2] | 0.9 [0.7-1.3] | 0.9 [0.6-1.1] |
| - Maximal neutrophils count (G/L) | 5.8 [4.0-7.9] | 5.7 [4.1-7.8] | 6.3 [4.3-8.0] | 5.7 [3.9-8.3] | 5.6 [3.8-7.8] |
| - Maximal plasma creatinine<br>(μmol/L) | 74.0 [62.0-91.0] | 72.5 [60.0-88.0] | 73.5 [62.0-88.0] | 77.0 [65.0-91.0] | 74.0 [62.0-93.0] |
| - Maximal SGOT (U/L) | 49.0 [35.0-72.0] | 53.0 [38.0-74.0] | 47.0 [34.0-64.0] | 47.0 [35.0-70.0] | 53.5 [34.0-81.0] |
| - Maximal SGPT (U/L) | 37.0 [25.0-63.0] | 41.0 [25.0-62.0] | 34.0 [22.5-60.5] | 37.0 [24.0-59.0] | 41.5 [26.0-67.0] |
| - Maximal plasma C-reactive protein<br>(mg/L) | 119.5 [72.0-185.0] | 132.0 [86.0-191.0] | 124.0 [75.0-188.0] | 105.0 [57.0-164.0] | 118.0 [72.0-188.0] |
| - Maximal plasma D-dimers (μg/L) | 1080.0 [649.0-1860.0] | 1170.0 [689.0-2000.0] | 1060.0 [626.0-1987.0] | 956.0 [560.0-1673.0] | 1140.0 [654.0-1820.0] |
| - Maximal procalcitonin (ng/mL) | 0.2 [0.1-0.9] | 0.3 [0.1-1.1] | 0.2 [0.1-0.6] | 0.3 [0.1-0.9] | 0.3 [0.1-0.9] |
| - Maximal ferritin (mg/L) | 480.5 [2.0-1344.0] | 98.0 [2.0-1041.0] | 608.0 [2.0-1288.0] | 761.0 [3.0-1344.0] | 377.0 [2.0-1610.0] |

### Primary endpoint

The distribution of the 7-point ordinal scale at day 15 is presented in the Figure 1 and Table 2. There was no significant difference between any of the treatment arm and the control arm, and adjusted OR (aOR) were not in favor of investigational treatments (*i*.*e*., below 1): lopinavir/ritonavir *versus* control, aOR 0.83 (95% confidence interval [CI] 0.55; 1.26, P=0.39); lopinavir/ritonavir plus IFN-ß-1a *vs*. control, aOR 0.69 (95%CI 0.45; 1.04, P=0.08); hydroxychloroquine *vs*. control, aOR 0.93 (95%CI 0.62; 1.41, P=0.75).

**Table 2.** Primary and secondary outcomes for patients included in the present analysis DisCoVeRy trial, according to disease severity at baseline. Analyses were stratified on the disease severity at baseline (moderate: 7-point ordinal scale 3 or 4; severe: 7-point ordinal scale 5 or 6), and adjusted effect measures are reported in the table. NP, Nasopharyngeal; LRT, Lower respiratory tract; OR, Odds-ratio; HR, Hazard ratio; LSMD, least-square mean difference.

| | Overall<br>(N=583) | | Control<br>(N=148) | | Lopinavir/ritonavir<br>(L/r)<br>(N=145) | | Lopinavir/ritonavir +<br>interferon β-1a<br>(L/r + IFN)<br>(N=145) | | Hydroxychloroquine<br>(HCQ)<br>(N=145) | | L/r<br>vs. control<br>Effect<br>measure<br>(95% CI) | L/r + IFN<br>vs. control<br>Effect<br>measure<br>(95% CI) | HCQ<br>vs. control<br>Effect<br>measure<br>(95% CI) |
| --- | --- | --- | --- | --- | --- | --- | --- | --- | --- | --- | --- | --- | --- |
|  | Moderate<br>(N=372) | Severe<br>(N=211) | Moderate<br>(N=94) | Severe<br>(N=54) | Moderate<br>(N=94) | Severe (N=51) | Moderate<br>(N=91) | Severe<br>(N=54) | Moderate<br>(N=93) | Severe<br>(N=52) |  |  |  |
| **7-point ordinal scale at day 15, n (%)** |  |  |  |  |  |  |  |  |  |  |  |  |  |
| 1. Not hospitalized, no limitations on activities | 84 (22.6%) | 3 (1.4%) | 23 (24.5%) | 1 (1.9%) | 21 (22.3%) | 1 (2.0%) | 20 (22.0%) | 0 (0.0%) | 20 (21.5%) | 1 (1.9%) | OR=0.83<br>(0.55 to 1.26)<br>[P=0.39] | OR=0.69<br>(0.45 to 1.04)<br>[P=0.08] | OR=0.93<br>(0.62 to 1.41)<br>[P=0.75] |
| 2. Not hospitalized, limitation on activities | 146 (39.2%) | 16 (7.6%) | 41 (43.6%) | 6 (11.1%) | 36 (38.3%) | 2 (3.9%) | 35 (38.5%) | 1 (1.9%) | 34 (36.6%) | 7 (13.5%) |  |  |  |
| 3. Hospitalized, not requiring supplemental oxygen | 54 (14.5%) | 22 (10.4%) | 7 (7.4%) | 5 (9.3%) | 16 (17.0%) | 5 (9.8%) | 13 (14.3%) | 5 (9.3%) | 18 (19.4%) | 7 (13.5%) |  |  |  |
| 4. Hospitalized, requiring supplemental oxygen | 41 (11.0%) | 31 (14.7%) | 12 (12.8%) | 10 (18.5%) | 9 (9.6%) | 9 (17.6%) | 9 (9.9%) | 6 (11.1%) | 11 (11.8%) | 6 (11.5%) |  |  |  |
| 5. Hospitalized, on non-invasive ventilation or high flow oxygen devices | 6 (1.6%) | 10 (4.7%) | 1 (1.1%) | 2 (3.7%) | 2 (2.1%) | 1 (2.0%) | 2 (2.2%) | 4 (7.4%) | 1 (1.1%) | 3 (5.8%) |  |  |  |
| 6. Hospitalized, on invasive mechanical ventilation or ECMO | 27 (7.3%) | 106 (50.2%) | 6 (6.4%) | 24 (44.4%) | 7 (7.4%) | 29 (56.9%) | 9 (9.9%) | 28 (51.9%) | 5 (5.4%) | 25 (48.1%) |  |  |  |
| 7. Death | 14 (3.8%) | 23 (10.9%) | 4 (4.3%) | 6 (11.1%) | 3 (3.2%) | 4 (7.8%) | 3 (3.3%) | 10 (18.5%) | 4 (4.3%) | 3 (5.8%) |  |  |  |
| **7-point ordinal scale at day 29, n (%)** |  |  |  |  |  |  |  |  |  |  |  |  |  |
| 1. Not hospitalized, no limitations on activities | 146 (39.2%) | 21 (10.0%) | 35 (37.2%) | 7 (13.0%) | 36 (38.3%) | 6 (11.8%) | 35 (38.5%) | 1 (1.9%) | 40 (43.0%) | 7 (13.5%) | OR=0.93<br>(0.62 to 1.41)<br>[P=0.74] | OR=0.76<br>(0.50 to 1.15)<br>[P=0.19] | OR=1.16<br>(0.77 to 1.75)<br>[P=0.49] |
| 2. Not hospitalized, limitation on activities | 128 (34.4%) | 29 (13.7%) | 35 (37.2%) | 5 (9.3%) | 36 (38.3%) | 6 (11.8%) | 29 (31.9%) | 8 (14.8%) | 28 (30.1%) | 10 (19.2%) |  |  |  |
| 3. Hospitalized, not requiring supplemental oxygen | 45 (12.1%) | 45 (21.3%) | 12 (12.8%) | 15 (27.8%) | 10 (10.6%) | 9 (17.6%) | 11 (12.1%) | 10 (18.5%) | 12 (12.9%) | 11 (21.2%) |  |  |  |
| 4. Hospitalized, requiring supplemental oxygen | 14 (3.8%) | 19 (9.0%) | 4 (4.3%) | 6 (11.1%) | 4 (4.3%) | 4 (7.8%) | 4 (4.4%) | 6 (11.1%) | 2 (2.2%) | 3 (5.8%) |  |  |  |
|  | Moderate<br>(N=372) | Severe<br>(N=211) | Moderate<br>(N=94) | Severe<br>(N=54) | Moderate<br>(N=94) | Severe (N=51) | Moderate<br>(N=91) | Severe<br>(N=54) | Moderate<br>(N=93) | Severe<br>(N=52) |  |  |  |
| 5. Hospitalized, on non-invasive ventilation or high flow oxygen devices | 5 (1.3%) | 10 (4.7%) | 2 (2.1%) | 1 (1.9%) | 1 (1.1%) | 2 (3.9%) | 1 (1.1%) | 3 (5.6%) | 1 (1.1%) | 4 (7.7%) |  |  |  |
| 6. Hospitalized, on invasive mechanical ventilation or ECMO | 14 (3.8%) | 52 (24.6%) | 1 (1.1%) | 13 (24.1%) | 3 (3.2%) | 14 (27.5%) | 6 (6.6%) | 13 (24.1%) | 4 (4.3%) | 12 (23.1%) |  |  |  |
| 7. Death | 20 (5.4%) | 35 (16.6%) | 5 (5.3%) | 7 (13.0%) | 4 (4.3%) | 10 (19.6%) | 5 (5.5%) | 13 (24.1%) | 6 (6.5%) | 5 (9.6%) |  |  |  |
| **Time to improvement of 2 categories of the 7-point ordinal scale or hospital discharge within day 29 (days), median [IQR]** | 10 [7-16] | 19 [14-29] | 9 [6-14] | 19 [10-29] | 11 [7-17] | 27 [14-29] | 10 [7-19] | 22 [15-29] | 10 [7-17] | 18 [13-29] | HR=0.71<br>(0.54 to 0.93)<br>[P=0.012] | HR=0.70<br>(0.54 to 0.92)<br>[P=0.009] | HR=0.79<br>(0.61 to 1.03)<br>[P=0.08] |
| **Time to National Early Warning Score $\leq 2$ or hospital discharge within 29 days (days), median [IQR]** | 9 [5-16] | 29 [17-29] | 8 [5-14] | 26 [15-29] | 9 [6-16] | 29 [22-29] | 9 [6-18] | 29 [19-29] | 9 [5-15] | 29 [16-29] | HR=0.83<br>(0.63 to 1.09)<br>[P=0.17] | HR=0.75<br>(0.56 to 0.99)<br>[P=0.046] | HR=0.90<br>(0.68 to 1.18)<br>[P=0.45] |
| **Time to hospital discharge within 29 days (days), median [IQR]** | 10 [7-20] | 29 [19-29] | 9 [6-16] | 29 [19-29] | 12 [8-21] | 29 [24-29] | 11 [8-26] | 29 [28-29] | 11 [7-20] | 29 [16-29] | HR=0.77<br>(0.58 to 1.02)<br>[P=0.07] | HR=0.72<br>(0.54 to 0.96)<br>[P=0.026] | HR=0.83<br>(0.62 to 1.10)<br>[P=0.20] |
| **Oxygenation-free days until day 29 (days), median [IQR]** | 22 [15-25] | 0 [0-13] | 22 [15-25] | 4 [0-14] | 22 [15-25] | 0 [0-12] | 22 [13-25] | 0 [0-6] | 22 [16-25] | 3 [0-15] | LSMD=-0.86<br>(-2.80 to 1.08)<br>[P=0.39] | LSMD=-1.68<br>(-3.66 to 0.29)<br>[P=0.10] | LSMD=0.17<br>(-1.84 to 2.17)<br>[P=0.87] |
| **Ventilator-free days until day 29 (days), median [IQR]** | 29 [29-29] | 11 [0-20] | 29 [29-29] | 14 [0-22] | 29 [29-29] | 3 [0-19] | 29 [29-29] | 4 [0-16] | 29 [29-29] | 14 [1-22] | LSMD=-0.98<br>(-2.96 to 1.00)<br>[P=0.33] | LSMD=-2.01<br>(-4.03 to 0.00)<br>[P=0.05] | LSMD=0.09<br>(-1.93 to 2.10)<br>[P=0.93] |
| **Death within 28 days, no. (%)** | 19 (5.1%) | 35 (16.6%) | 5 (5.3%) | 7 (13.0%) | 4 (4.3%) | 10 (19.6%) | 4 (4.4%) | 13 (24.1%) | 6 (6.5%) | 5 (9.6%) | OR=1.24<br>(0.55 to 2.82)<br>[P=0.60] | OR=1.51<br>(0.69 to 3.34)<br>[P=0.30] | OR=0.93<br>(0.40 to 2.20)<br>[P=0.88] |

**Figure 1.**
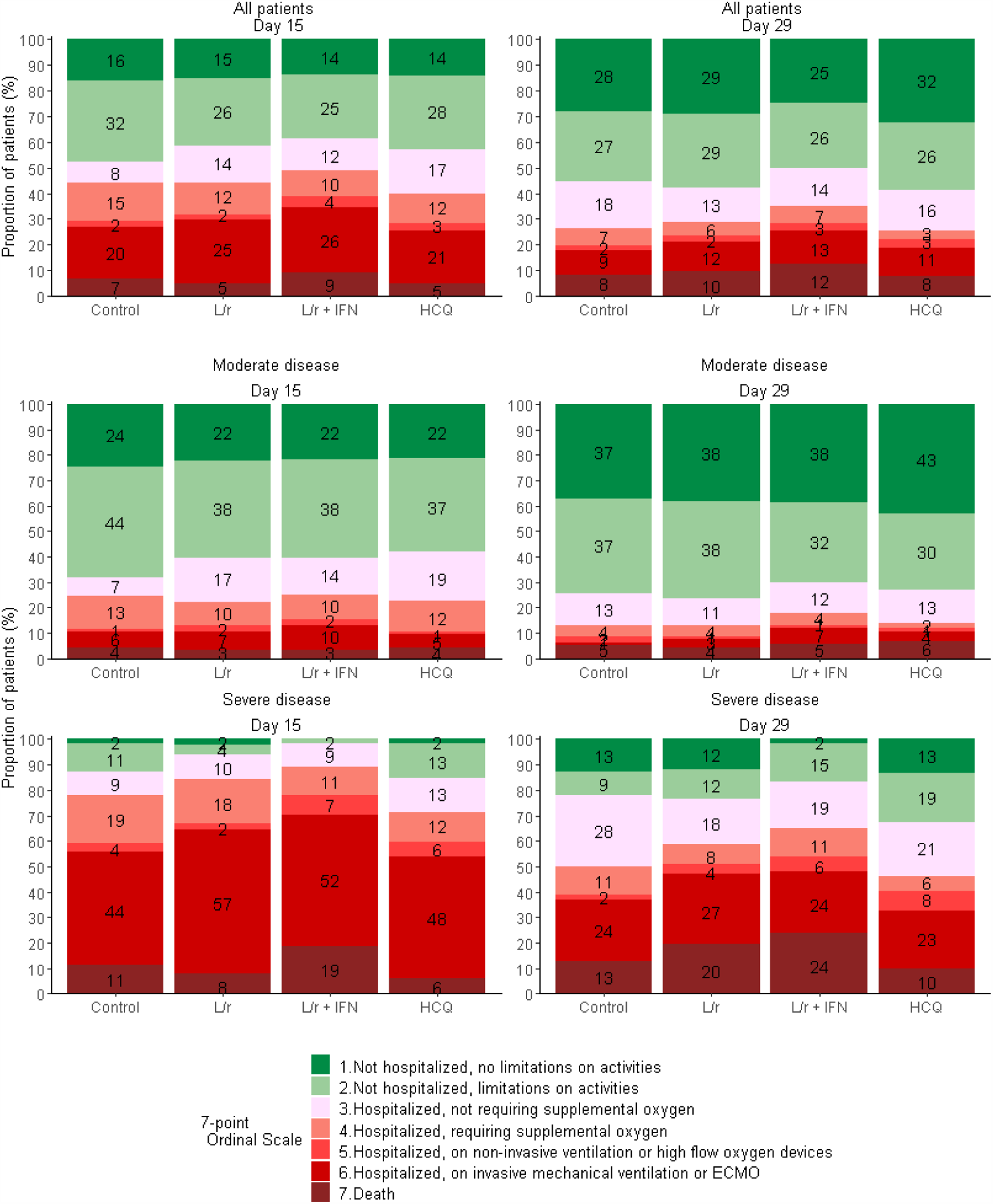
Clinical status, as measured by the 7-point ordinal scale, at day 15 and day 29 of patients from the intention-to-treat population of the DisCoVeRy trial, according to treatment arm and disease severity at baseline. Reported numbers refer to the proportion of patients with the corresponding level in each group. L/r, Lopinavir/ritonavir; L/r + IFN, Lopinavir/ritonavir + interferon ß-1a; HCQ, Hydroxychloroquine.

### Secondary endpoints

The distribution of the 7-point ordinal scale at day 29 is presented in the Figure 1 and Table 2. There was no significant difference between any of the treatment arm and the control arm. The time to improvement of 2 categories of the 7-point ordinal scale or hospital discharge within day 29 was significantly higher in both lopinavir/ritonavir-containing arms than in the control arm. The time to NEWS ≤2 or hospital discharge within 29 days and time to hospital discharge within day 29 were significantly higher in the lopinavir/ritonavir plus IFN-ß-1a arm than in the control arm (Table 2). No other significant difference was observed for any other secondary outcomes between treatment arms and the control arm (Table 2 and Supplementary Figures S2-S4).

### Virological endpoints

No significant difference in the proportion of participants with detectable log_10_ normalized viral loads in NPS at each sampling time was demonstrated (Supplementary Table S2). The proportion of participants having severe disease with detectable log_10_ normalized viral loads in the LRT at each sampling time is presented in Supplementary Table S3.

The slope of decrease of the log_10_ normalized viral loads quantified in NPS over time was not significantly affected by any of the investigational treatment (Figure 2, panel A).

**Figure 2.**
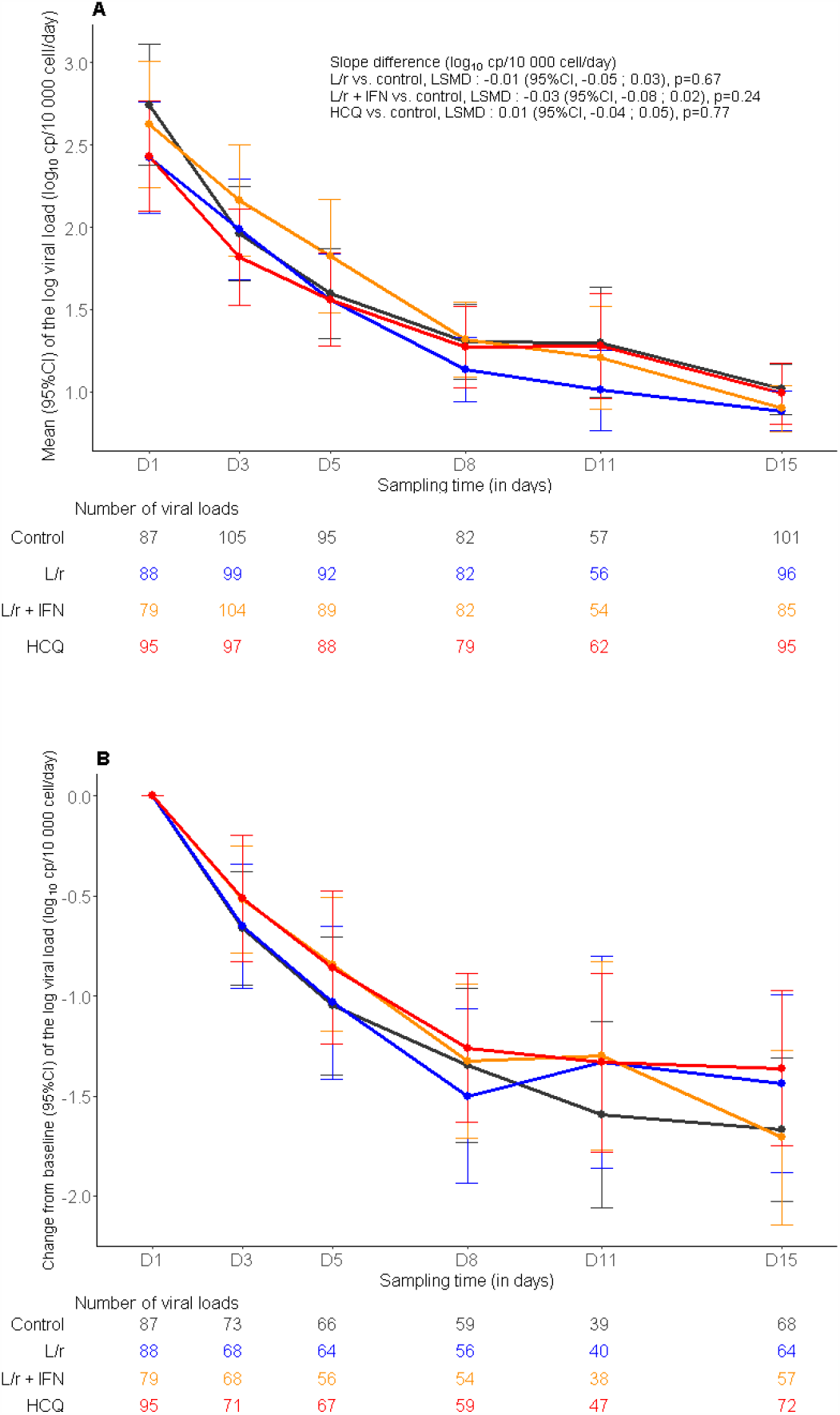
Evolution of the normalized SARS-CoV-2 viral load in nasopharyngeal swabs between baseline and day 15 in the intention-to-treat population of the DisCoVeRy trial: means (95%CI) of the log viral loads (panel A), mean changes from baseline (95%CI) of the log viral loads (panel B). L/r, Lopinavir/ritonavir (blue line); L/r + IFN, Lopinavir/ritonavir + interferon ß-1a (yellow line); HCQ, Hydroxychloroquine (red line); control (black line). LSMD, least-square mean difference; 95%CI, 95% confidence interval.

### Safety

A total of 579 participants were included in the safety analysis (control, n=148; lopinavir/ritonavir, n=144; lopinavir/ritonavir plus IFN-β-1a, n=144; hydroxychloroquine, n=143). Safety outcomes are presented in the Table 3. Among the 2399 reported AEs, 477 were graded 3 or 4 in 205 patients and mostly reported in the lopinavir/ritonavir arms (Table 3).

Consistently, 608 serious adverse events (SAEs) were reported in 274 participants; 149 (24.5%) were considered to be related to the investigational drug according to investigator’s judgment (accounting for 37 events in the lopinavir/ritonavir arm, 71 events in the lopinavir/ritonavir plus IFN-β-1a arm, and 41 events in the hydroxychloroquine arm). A significantly higher number of patients experienced at least one SAE in the lopinavir/ritonavir-containing arms than in the control arm (76/144, 52.8% in the lopinavir/ritonavir arm *vs*. 57/148, 38.5% in the control arm, p=0.02; 78/144, 54.2% in the lopinavir/ritonavir plus IFN-ß-1a arm *vs*. 57/148, 38.5% in the control arm, p=0.01).

**Table 3.** Summary of adverse events according treatment group in the modified intention to treat population. In the “Overall” column, numbers refer to number of events and number of patients. In other columns, number refer to number of patients (%). Some patients had more than a single SAE. Analyses were performed on the modified Intention-to-treat population. SAE, Serious Adverse Event. P-value refer to Fisher exact test. * Including renal failure in 30 patients, hepatic disorders in 18 patients and electrocardiogram abnormalities in 8 patients ** Excluding acute renal failures defined based on the RIFLE classification.

|  | **Overall**<br>**(N=579)** | **Control**<br>**(N=148)** | **Lopinavir/ritonavir**<br>**(L/r)**<br>**(N=144)** | **Lopinavir/ritonavir +**<br>**Interferon β-1a**<br>**(L/r + IFN)**<br>**(N=144)** | **Hydroxychloroquine**<br>**(HCQ)**<br>**(N=143)** | **L/r**<br>**vs. control**<br>**P-value** | **L/r + IFN**<br>**vs. control**<br>**P-value** | **HCQ**<br>**vs. control**<br>**P-value** |
| --- | --- | --- | --- | --- | --- | --- | --- | --- |
|  | **no. events /**<br>**no. patients** | **no.**<br>**patients**<br>**(%)** | **no. patients (%)** | **no. patients (%)** | **no. patients(%)** |  |  |  |
| **Any adverse events** | 2399 / 450 | 105<br>(70.9%) | 119 (82.6%) | 117 (81.3%) | 109 (76.2%) | 0.02 | 0.04 | 0.35 |
| **Any grade 3 or 4 adverse events** | 477 / 205 | 48 (32.4%) | 56 (38.9%) | 58 (40.3%) | 43 (30.1%) | 0.27 | 0.18 | 0.71 |
| **Any serious adverse events** | 608 / 274 | 57 (38.5%) | 76 (52.8%) | 78 (54.2%) | 63 (44.1%) | 0.02 | 0.01 | 0.34 |
| **Premature suspension or**<br>**discontinuation of the**<br>**experimental treatment\*** | 77 (13.3%) | - | 17 (11.8%) | 43 (29.9%) | 17 (11.9%) | - | - | - |
| **Most relevant SAEs** |  |  |  |  |  |  |  |  |
| - Acute respiratory failure | 65 / 65 | 18 (12%) | 19 (13%) | 17 (12%) | 11 (8%) |  |  |  |
| - Acute Respiratory Distress<br>syndrome | 47 / 46 | 16 (11%) | 7 (5%) | 10 (7%) | 13 (9%) |  |  |  |
| - Acute kidney injury** | 50 / 50 | 9 (6%) | 16 (11%) | 11 (8%) | 14 (10%) |  |  |  |
| - Acute renal failure based on the<br>RIFLE classification | 17 / 17 | 3 (2%) | 3 (2%) | 8 (6%) | 3 (2%) |  |  |  |
| - Arrhythmia | 41 / 35 | 3 (2%) | 8 (6%) | 12 (8%) | 12 (8%) |  |  |  |
| - Pulmonary embolism | 27 / 27 | 6 (4%) | 10 (7%) | 5 (3%) | 6 (4%) |  |  |  |
| - Transaminases increased | 25 / 25 | 2 (1%) | 5 (3%) | 12 (8%) | 6 (4%) |  |  |  |
| - Sepsis | 21 / 21 | 2 (1%) | 6 (4%) | 7 (5%) | 6 (4%) |  |  |  |
| - Cholestasis | 6 / 6 | 0 (0%) | 2 (1%) | 3 (2%) | 1 (1%) |  |  |  |

The most frequently reported SAEs were acute respiratory failure (n=65, 11%), acute kidney injury (n=50, 8.2%), acute respiratory distress syndrome (n=47, 8%), arrhythmia (n=41, 7%), pulmonary embolism (n=27, 5%), and sepsis including those related to super-infections (n=21, 4%). Thirteen percent (n=76) of participants developed at least one kidney-related SAE; 66 of them were critically ill ventilated patients associated with acute kidney injury or acute renal failure, of whom 12 had acute renal failure at baseline. Among the 57 fatal SAEs, 23 were classified as having a pulmonary origin, and 34 as having a non-pulmonary origin. Only 4 non-pulmonary-related deaths were linked to investigational treatments by the investigators (lopinavir/ritonavir arm, n=1; lopinavir/ritonavir plus IFN-ß-1a arm, n=3).

## Discussion

We report here the results of the DisCoVeRy clinical trial evaluating lopinavir/ritonavir with or without IFN-ß-1a, or hydroxychloroquine in comparison with control for the treatment of inpatients with COVID-19. As reported by the Solidarity interim analysis of critical endpoints, we report that these investigational treatments failed to improve the clinical course of COVID-19, nor to enhance SARS-CoV-2 clearance and were associated with more SAEs. Several larger-scale randomized controlled trials conducted in hospitalized COVID-19 patients failed to demonstrate the efficacy of hydroxychloroquine either by addressing the question through the improvement in the 7-point ordinal scale at 15 days(20) or through the effect on 28-day mortality(21) as compared to SoC alone. Our results are in line with these conclusions. We bring additional evidence that hydroxychloroquine does not accelerate SARS-CoV-2 RNA clearance, consistently with preclinical data in a macaque model(22).

Two randomized trials conducted in hospitalized COVID-19 patients found no difference between the lopinavir/ritonavir and the SoC arms in terms of 28-day mortality or of probability of progression to mechanical ventilation or death(23, 24). The hypothesis that severe COVID-19 is associated with an impaired type I IFN response characterized by no or low IFN production and activity supported testing IFN with lopinavir/ritonavir. In an open-label randomized trial, the combination of lopinavir/ritonavir, IFN-β-1b and ribavirin was superior to lopinavir/ritonavir in alleviating symptoms and shortening the duration of SARS-CoV-2 shedding in mild-to-moderate COVID-19(25). Conversely, our data suggest longer time to NEW2 improvement or hospital discharge in the lopinavir/ritonavir plus IFN-β-1a *vs*. lopinavir/ritonavir arm as well as no virological effect (Supplementary Figure S3). In addition, we observed a higher rate of SAEs in the lopinavir/ritonavir-containing arms. Participants from the investigational arms experienced more acute kidney injury, although the difference was not significant. In the first series of COVID-19, acute renal failure incidence ranged from 3 to 25% among heterogeneous patients in terms of disease severity and treatment management, although predominantly in critically ill patients(26, 27). COVID-19 has been associated with renal endotheliitis(28). Elevation of serum creatinine has been associated with high levels of D-dimers (> 500 ng/mL)(29), which resonates with elevated median D-dimer levels at baseline. As reported with HIV-infected patients, infection and inflammation are associated with down-regulation of cytochrome P450(30, 31). Gregoire et al reported that intense systemic inflammatory response to SARS-CoV-2 infection enables important increase in lopinavir unbound plasma concentrations(32). Marzolini et al found that lopinavir concentrations were significantly correlated with a CRP value above 75 mg/L(33), most of our participants being above this value at baseline. Lê et al demonstrated that in mechanically-ventilated patients with COVID-19, the oral administration of lopinavir/ritonavir elicited plasma exposure of lopinavir > 6-fold the upper usual expected range(34), consistent with the fact that all kidney-related SAE occurred in critically ill ventilated patients.

DisCoVeRy is an adaptive, randomized trial initiated in France and that has been now extended to other European countries with the support of the European commission and the task of becoming a pan-European platform trial for Emerging infectious Diseases treatment. Furthermore, DisCoVeRy is an add-on trial to Solidarity and importantly contributes to Solidarity data acquisition and early interruption of treatment arms for futility. In agreement with Solidarity, our objective was to complement knowledge acquisition with granular safety and virological data. The design and the philosophy of DisCoVeRy trial will be of even greater importance when new investigational drugs will be evaluated, for which scarce safety data might be available, unlike repurposed drugs. The trial has some limitations: it is not a placebo-controlled, double-blinded trial due to the complexity of blinding treatments with different mode of administration (intravenous, subcutaneous or oral) and the need to initiate the trial very rapidly. DisCoVeRy did not target patients at the early phase of the disease nor include arms testing anti-inflammatory agents that can be used as part of the standard of care in any arm.

## Conclusion

In patients admitted to hospital with COVID-19, lopinavir/ritonavir, lopinavir/ritonavir plus IFN-β-1a and hydroxychloroquine were not associated with clinical improvement at day 15 and day 29, nor reduction in viral shedding, and generated substantial SAEs. These findings do not support the use of these investigational treatments for patients hospitalized with COVID-19.

## Data Availability

Data will be made available upon reasonnable request to the corresponding author

## Notes

### Competing Interest Statement

The authors have declared no competing interest.

### Clinical Trial

NCT04315948

### Author Declarations

The trial was approved by the Ethics Committee (CPP Ile-de-France-III, approval #20.03.06.51744).

