## Supplementary Appendix for "Antiviral drugs in hospitalized patients with COVID-19 - the DisCoVeRy trial"

by Ader, F *et al.*

### Supplementary Appendix

#### Supplementary Methods

##### *Selection criteria*

We enrolled adult patients who were hospitalized with PCR-confirmed COVID-19, as determined by a positive SARS-CoV-2 RT-PCR on any biological sample.

The full list of inclusion and exclusion criteria was the following.

Inclusion criteria:

- Adult ≥18 years of age at time of enrolment;
- laboratory-confirmed SARS-CoV-2 infection as determined by PCR, or other commercial or public health assay in any specimen < 72 hours prior to randomization;
- Hospitalized patients with illness of any duration, and at least one of the following:
  - Clinical assessment (evidence of rales/crackles on physical examination) AND SpO2 ≤ 94% on room air, OR
  - Acute respiratory failure requiring supplemental oxygen, high flow oxygen devices, non-invasive ventilation, and/or mechanical ventilation;
- Women of childbearing potential must agree to use contraception for the duration of the study.

Exclusion criteria:

- Refusal to participate expressed by patient or legally authorized representative if they are present;
- Spontaneous blood alanine transferase (ALT)/AST levels > 5 times the upper limit of normal;

- Stage 4 severe chronic kidney disease or requiring dialysis (i.e. eGFR < 30 mL/min);
- Pregnancy or breast-feeding;
- Anticipated transfer to another hospital, which is not a study site within 72 hours;
- Patients previously treated with one of the antivirals evaluated in the trial (i.e. remdesivir, interferon ß-1a, lopinavir/ritonavir, hydroxychloroquine) in the past 29 days;
- Contraindication to any study medication including allergy;
- Use of medications that are contraindicated with lopinavir/ritonavir i.e. drugs whose metabolism is highly dependent on the isoform CYP3A with narrow therapeutic range (e.g. amiodarone, colchicine, simvastatine);
- Use of medications that are contraindicated with hydroxychloroquine: citalopram, escitalopram, hydroxyzine, domperidone, pipéraquine;
- Human immunodeficiency virus infection under highly active antiretroviral therapy (HAART);
- History of severe depression or attempted suicide or current suicidal ideation;
- Corrected QT interval superior to 500 milliseconds (as calculated with the Fridericia formula).

***Sample size calculation***

Hypotheses for sample size calculation are presented in Supplementary Material. The sample size was determined assuming the following scenario under standard of care for each item of the ordinal scale at day 15: 1, 42%; 2, 38%; 3, 8%; 4, 7%; 5, 2%; 6, 1%; 7, 2%. At the time of the trial design (March 2020), there was a significant uncertainty with these assumptions. We powered the study for an odds ratio of 1.5 (an odds ratio higher than 1 indicates superiority of the experimental treatment over the control for each ordinal scale category), with 90% power and using an overall one-sided type I error rate of 0.05. Adjusting for multiplicity of 4 pairwise comparisons with the control arm in a 5-arm setting, the (one-sided) false positive error rate would be 0.00625. We determined that the inclusion of 620 patients in each treatment arm was required.

***Virological Methods***

Nasopharyngeal swabs were collected by trained practitioners using validated devices containing flocked swabs and virus transport medium. Low respiratory samples were collected in sterile containers and were sent, as swabs, to the laboratory to be processed within 24 hours after sampling for SARS-CoV-2 detection. Each sample was then sent after freezing, to the National Center for Viral Respiratory Infections (Hospices Civils de Lyon, France) for the determination of the normalized viral load. The analysis of the swab was performed blinded to treatment arm. RNA extraction was performed on the EMAG® platform (bioMerieux, Marcy-l'Étoile, France), using manufacturer's instructions. RNA was extracted from 200 µl of sample eluted in 50 µl of elution buffer. Then, SARS-CoV-2 viral load was measured by quantitative RT-PCR using the RT-PCR RdRp-IP4 developed by the French National Center for Viral Respiratory Infections (Institut Pasteur, Paris) widely used in French laboratories at the beginning of the pandemic[33]. The amplification protocol was developed for viral quantification using QuantStudio 5 *rt*PCR Systems (Thermo Fisher Scientific, Waltham, Massachusetts, USA). This RT-PCR is targeting RdRp gene and is one of the most sensitive method available[33]. SARS-COV-2 viral loads were measured according a scale of calibrated in-house plasmid, ranging from $5.10^2$ to $5.10^6$ copies/5 µL. The quality of nasopharyngeal swabs was checked using the CELL Control r-gene® kit (Argene_BioMérieux, Marcy-l'Étoile, France). This kit is provided with a quantified plasmid for cellular quantification. If cell quantification was below 500 cells/5µL, we considered that the sample had too poor quality for viral load to be measured. We computed a normalized SARS-CoV-2 viral load by dividing the viral load measured in the sample by the number of cells measured, and expressed it in log10 of RNA copies per 10 000 cells. The limit of detection (LoD) of RT-PCR for the viral load was 4 copies / reaction, which corresponds to a limit of detection for normalized viral load of 1 log10 copies/100 000 cells. We estimated the LoD using a probit analysis by analyzing dilutions of a quantified cell culture supernatants. Probit analysis consists of describing the relationship between the probability of detection and concentration using a cumulative probability curve. For each dilution, the ratio (the hit rate) is computed as the number of replicates with a detected outcome per the total number of replicates tested. These hit rates are converted mathematically into cumulative normal probability units (probits) and fitted using a regression model vs. their respective concentrations. The LoD is defined as the lowest amount of viral genome that can be detected with a 95% hit rate. In the study, all viral load strictly under 1 was considered under the LoD and was reported in the e-CRF as a negative result. Any point of kinetics corresponding to a rebound of viral excretion was retested for confirmation.

***Handling of missing data***

For the 7-point ordinal scale, missing data were imputed using the last observation carried forward method, excepted in the case of known death or hospital discharge, in which case the ordinal scale was imputed to the value of 7 (death) or 2 (not hospitalized, limitation of activities), respectively. For NEWS, oxygenation and mechanical ventilation outcomes, missing data were treated using the last observation carried forward method, except in the case of preceding death, in which case patients were imputed to the worst value of the NEWS, or considered to have oxygen or require mechanical ventilation. For time-to-event analyses, patients were censored at day 29, at their date of loss of follow-up, or of study withdrawal, whichever occurred first. Deaths were censored at day 29. Missing SARS-CoV-2 viral loads were not imputed. For the analysis of viral load by mixed models, undetectable viral load values (i.e. values < 1 log10 copies/10 000 cells) were imputed to half the LoD hence 0.7 log10 copies/10 000 cells. In case of several consecutive undetectable values, only the first one was replaced and the subsequent discarded (until the next detectable value, if values were available afterwards).

#### Supplementary Figures

##### Supplementary Figure S1. Enrollment and randomization of patients in the present analysis of the DisCoVeRy trial.

Patients from all groups received the standard of care, in addition to the treatment allocated by randomization.

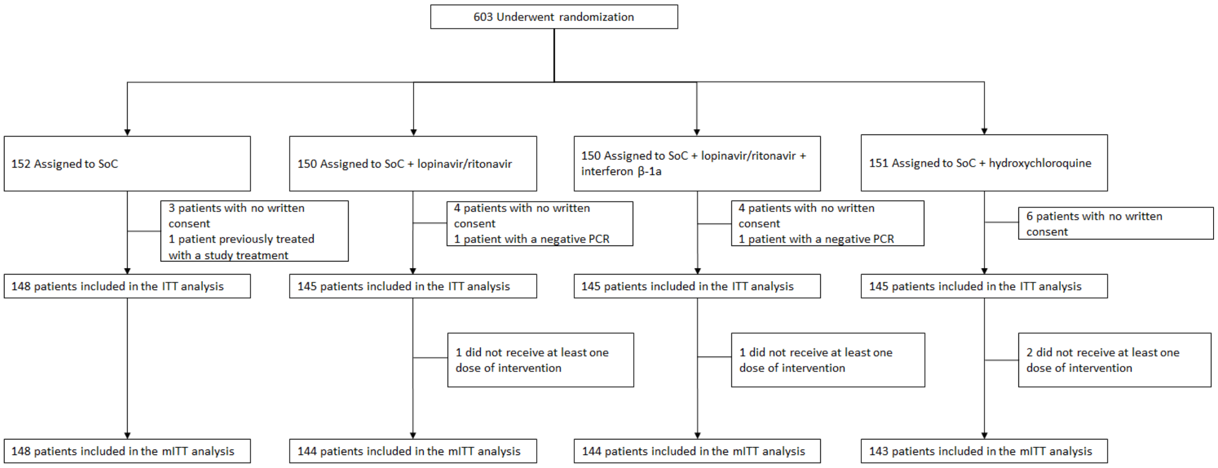

**Supplementary Figure S2. Time to improvement of at least 2 categories or the 7-point ordinal scale or hospital discharge between baseline and day 29 in the intention-to-treat population of the DisCoVeRy trial, according to disease severity at baseline in all participants (panel A), in participants with moderate disease at baseline (panel B) and in participants with severe disease at baseline (panel C).**

L/r, Lopinavir/ritonavir; L/r + IFN, Lopinavir/ritonavir + interferon ß-1a; HCQ, Hydroxychloroquine; HR, hazard ratio.

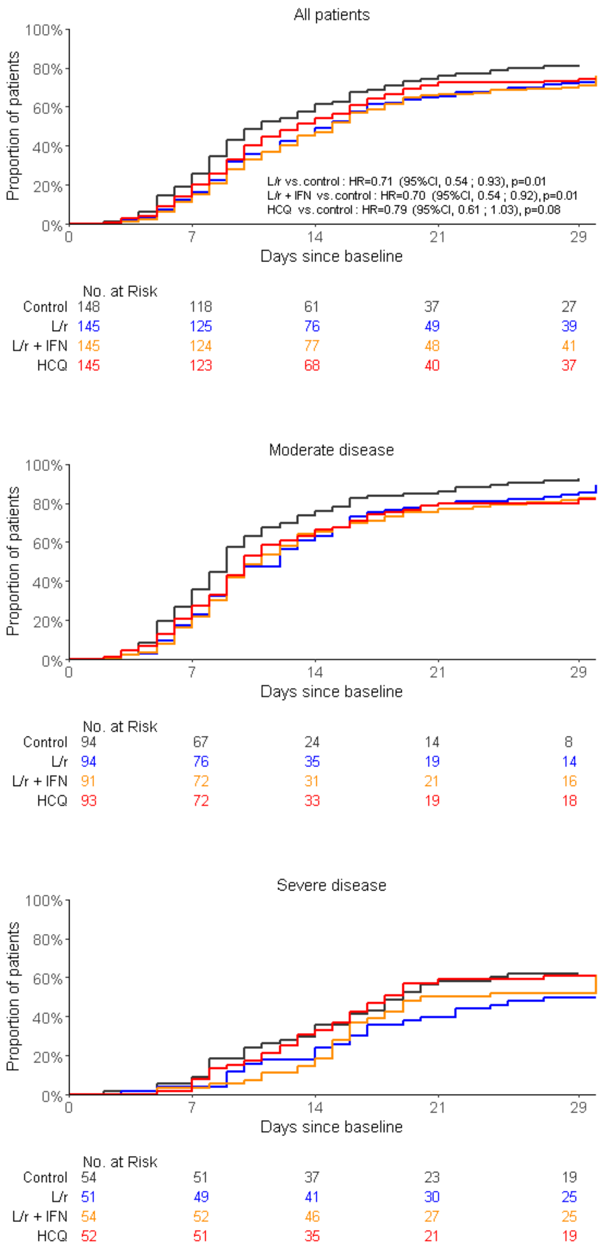

**Supplementary Figure S3. Time to National Early Warning Score ≤2 or hospital discharge between baseline and day 29 in the intention-to-treat population of the DisCoVeRy trial, according to disease severity at baseline in all participants (panel A), in participants with moderate disease at baseline (panel B) and in participants with severe disease at baseline (panel C).**

L/r, Lopinavir/ritonavir; L/r + IFN, Lopinavir/ritonavir + interferon ß-1a; HCQ, Hydroxychloroquine; HR, hazard ratio.

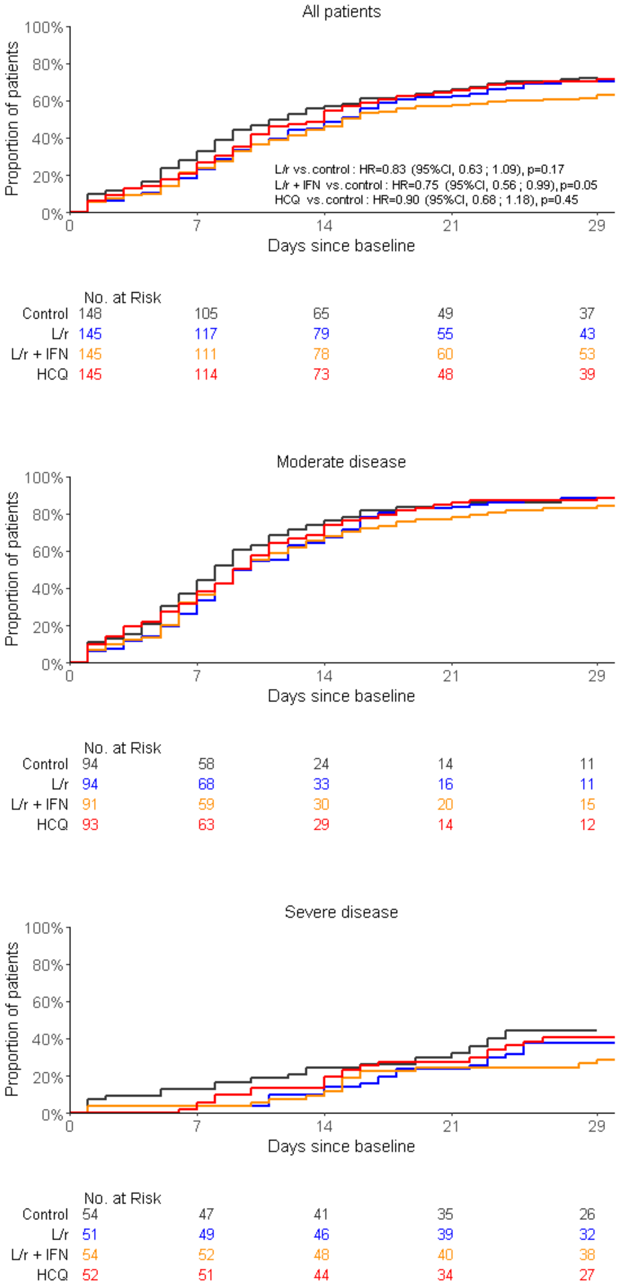

**Supplementary Figure S4. Time to hospital discharge within day 29 between baseline and day 29 in the intention-to-treat population of the DisCoVeRy trial, according to disease severity at baseline in all participants (panel A), in participants with moderate disease at baseline (panel B) and in participants with severe disease at baseline (panel C).**

L/r, Lopinavir/ritonavir; L/r + IFN, Lopinavir/ritonavir + interferon ß-1a; HCQ, Hydroxychloroquine; HR, hazard ratio.

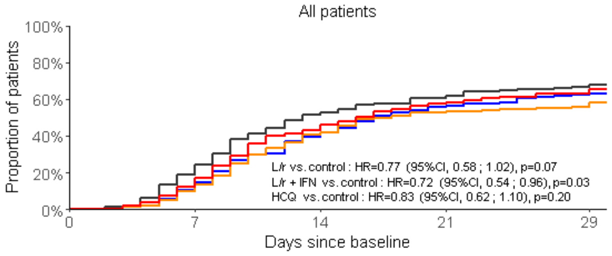


| No. at Risk | | | | | |
|---|---|---|---|---|---|
| Control | 148 | 120 | 71 | 57 | 47 |
| L/r | 145 | 130 | 87 | 64 | 54 |
| L/r + IFN | 145 | 130 | 85 | 68 | 62 |
| HCQ | 145 | 127 | 82 | 58 | 50 |

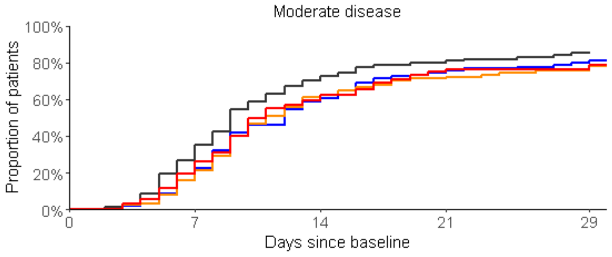


| No. at Risk | | | | | |
|---|---|---|---|---|---|
| Control | 94 | 69 | 28 | 19 | 14 |
| L/r | 94 | 79 | 39 | 24 | 19 |
| L/r + IFN | 91 | 76 | 35 | 26 | 22 |
| HCQ | 93 | 75 | 38 | 23 | 22 |

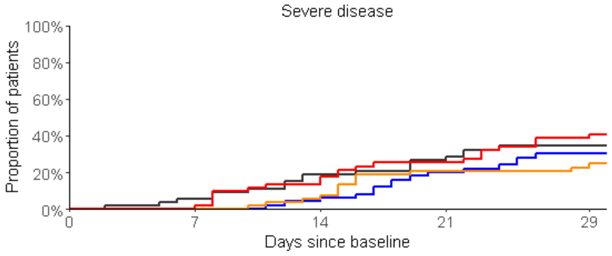


| No. at Risk | | | | | |
|---|---|---|---|---|---|
| Control | 54 | 51 | 43 | 38 | 33 |
| L/r | 51 | 51 | 48 | 40 | 35 |
| L/r + IFN | 54 | 54 | 50 | 42 | 40 |
| HCQ | 52 | 52 | 44 | 35 | 28 |

#### Supplementary Tables

##### Supplementary Table S1. Treatments received during the study course in the patients included in the intention-to-treat population of the DisCoVeRy trial.

| Treatment — no. (%) | Overall (N=583) | Control (N=148) | Lopinavir/ritonavir (L/r) (N=145) | Lopinavir/ritonavir + interferon ß-1a (L/r + IFN) (N=145) | Hydroxychloroquine (HCQ) (N=145) |
|---|---|---|---|---|---|
| **Corticosteroids** | 172 (29.5%) | 41 (27.7%) | 41 (28.3%) | 47 (32.4%) | 43 (29.7%) |
| - Dexamethasone | 42 (7.2%) | 7 (4.7%) | 9 (6.2%) | 14 (9.7%) | 12 (8.3%) |
| - Hydrocortisone | 35 (6.0%) | 10 (6.8%) | 7 (4.8%) | 12 (8.3%) | 6 (4.1%) |
| - Methyprednisolone | 56 (9.6%) | 14 (9.5%) | 14 (9.7%) | 15 (10.3%) | 13 (9.0%) |
| **Interleukin-6 inhibitors** | 6 (1.0%) | 3 (2.0%) | 1 (0.7%) | 1 (0.7%) | 1 (0.7%) |
| - Tocilizumab | 6 (1.0%) | 3 (2.0%) | 1 (0.7%) | 1 (0.7%) | 1 (0.7%) |
| - Sarilumab | 0 (0.0%) | 0 (0.0%) | 0 (0.0%) | 0 (0.0%) | 0 (0.0%) |
| **Interleukin-1 inhibitors** | 4 (0.7%) | 1 (0.7%) | 0 (0.0%) | 1 (0.7%) | 2 (1.4%) |
| **Antibiotics** | 495 (84.9%) | 131 (88.5%) | 128 (88.3%) | 117 (80.7%) | 119 (82.1%) |
| - Azithromycine | 38 (6.5%) | 8 (5.4%) | 9 (6.2%) | 10 (6.9%) | 11 (7.6%) |
| **Anticoagulants** | 549 (94.2%) | 138 (93.2%) | 136 (93.8%) | 137 (94.5%) | 138 (95.2%) |
| **Parenteral or enteral nutrition** | 198 (34.0%) | 49 (33.1%) | 52 (35.9%) | 53 (36.6%) | 44 (30.3%) |
| **Vasopressors** | 218 (37.4%) | 53 (35.8%) | 54 (37.2%) | 61 (42.1%) | 50 (34.5%) |
| **Extra-renal replacement/hemofiltration** | 54 (9.3%) | 8 (5.4%) | 17 (11.7%) | 21 (14.5%) | 8 (5.5%) |
| **Neuromuscular blocking agents** | 194 (33.3%) | 47 (31.8%) | 43 (29.7%) | 58 (40.0%) | 46 (31.7%) |
| **Inhaled nitric oxide** | 41 (7.0%) | 8 (5.4%) | 8 (5.5%) | 16 (11.0%) | 9 (6.2%) |
| **Prone positioning** | 187 (32.1%) | 37 (25.0%) | 43 (29.7%) | 66 (45.5%) | 41 (28.3%) |

#### Supplementary Table S2. Proportion of patients with detectable viral loads in the nasopharyngeal swabs at each sampling time, in the intention-to-treat population of the DisCoVeRy trial.

NPS, Nasopharyngeal swab.

| | **Overall (N=583)** | | **Control (N=148)** | | **Lopinavir/ritonavir (L/r) (N=145)** | | **Lopinavir/ritonavir + interferon ß-1a (L/r + IFN) (N=145)** | | **Hydroxychloroquine (HCQ) (N=145)** | | **L/r vs. control Effect measure (95%CI)** | **L/r + IFN vs. control Effect measure (95%CI)** | **HCQ vs. control Effect measure (95%CI)** |
|---|---|---|---|---|---|---|---|---|---|---|---|---|---|
| | **Moderate (N=372)** | **Severe (N=211)** | **Moderate (N=94)** | **Severe (N=54)** | **Moderate (N=94)** | **Severe (N=51)** | **Moderate (N=91)** | **Severe (N=54)** | **Moderate (N=93)** | **Severe (N=52)** | | | |
| **Detectable viral load in NPS**, n/N (%) | 144/265 (54.3%) | 79/128 (61.7%) | 33/66 (50.0%) | 24/33 (72.7%) | 36/66 (54.5%) | 19/30 (63.3%) | 39/68 (57.4%) | 20/33 (60.6%) | 36/65 (55.4%) | 16/32 (50.0%) | OR=1.00 (0.57 to 1.78) [P=0.99] | OR=1.04 (0.59 to 1.83) [P=0.89] | OR=0.85 (0.49 to 1.50) [P=0.58] |
| **Detectable viral load in NPS at day 5**, n/N (%) | 100/245 (40.8%) | 57/114 (50.0%) | 21/63 (33.3%) | 18/31 (58.1%) | 23/63 (36.5%) | 13/28 (46.4%) | 30/60 (50.0%) | 15/27 (55.6%) | 26/59 (44.1%) | 11/28 (39.3%) | OR=0.94 (0.52 to 1.70) [P=0.83] | OR=1.54 (0.85 to 2.78) [P=0.15] | OR=1.05 (0.58 to 1.88) [P=0.88] |
| **Detectable viral load in NPS at day 8**, n/N (%) | 60/199 (30.2%) | 44/122 (36.1%) | 17/50 (34.0%) | 10/30 (33.3%) | 14/50 (28.0%) | 7/31 (22.6%) | 17/47 (36.2%) | 14/34 (41.2%) | 12/52 (23.1%) | 13/27 (48.1%) | OR=0.69 (0.35 to 1.36) [P=0.28] | OR=1.21 (0.64 to 2.31) [P=0.56] | OR=0.93 (0.48 to 1.79) [P=0.82] |
| **Detectable viral load in NPS at day 11,** n/N (%) | 23/120 (19.2%) | 29/102 (28.4%) | 6/29 (20.7%) | 6/25 (24.0%) | 4/31 (12.9%) | 6/25 (24.0%) | 4/26 (15.4%) | 10/26 (38.5%) | 9/34 (26.5%) | 7/26 (26.9%) | OR=0.77 (0.30 to 1.96) [P=0.58] | OR=1.26 (0.52 to 3.08) [P=0.61] | OR=1.28 (0.54 to 3.02) [P=0.58] |
| **Detectable viral load in NPS at day 15**, n/N (%) | 36/263 (13.7%) | 23/115 (20.0%) | 11/69 (15.9%) | 8/32 (25.0%) | 7/71 (9.9%) | 5/26 (19.2%) | 9/60 (15.0%) | 2/24 (8.3%) | 9/63 (14.3%) | 8/33 (24.2%) | OR=0.63 (0.28 to 1.38) [P=0.25] | OR=0.65 (0.29 to 1.46) [P=0.30] | OR=0.91 (0.44 to 1.89) [P=0.81] |
| **Detectable viral load in NPS at day 29**, n/N (%) | 10/231 (4.3%) | 4/88 (4.5%) | 3/59 (5.1%) | 2/19 (10.5%) | 3/63 (4.8%) | 1/21 (4.8%) | 2/54 (3.7%) | 0/23 (0.0%) | 2/55 (3.6%) | 1/25 (4.0%) | OR=0.73 (0.19 to 2.81) [P=0.65] | OR=0.40 (0.08 to 2.06) [P=0.25] | OR=0.55 (0.13 to 2.38) [P=0.42] |

**Supplementary Table S3. Number of severe patients with detectable normalized viral load in the lower respiratory tract at each sampling time, in the intention-to-treat population of the DisCoVeRy trial.**

| | **Overall (N=211)** | **Control (N=54)** | **Lopinavir/ritonavir (L/r) (N=51)** | **Lopinavir/ritonavir + interferon (L/r + IFN) (N=54)** | **Hydroxychloroquine (HCQ) (N=52)** |
|---|---|---|---|---|---|
| **Detectable viral load in LRT samples at day 3,** n/N (%) | 33/35 (94.3%) | 9/9 (100.0%) | 9/10 (90.0%) | 10/11 (90.9%) | 5/5 (100.0%) |
| **Detectable viral load in LRT samples at day 5,** n/N (%) | 36/39 (92.3%) | 7/7 (100.0%) | 10/10 (100.0%) | 11/11 (100.0%) | 8/11 (72.7%) |
| **Detectable viral load in LRT samples at day 8,** n/N (%) | 22/30 (73.3%) | 5/7 (71.4%) | 7/9 (77.8%) | 6/7 (85.7%) | 4/7 (57.1%) |
| **Detectable viral load in LRT samples at day 11**, n/N (%) | 15/26 (57.7%) | 5/8 (62.5%) | 5/6 (83.3%) | 3/7 (42.9%) | 2/5 (40.0%) |
| **Detectable viral load in LRT samples at day 15,** n/N (%) | 1/17 (5.9%) | 0/3 (0.0%) | 0/5 (0.0%) | 1/7 (14.3%) | 0/2 (0.0%) |
| **Detectable viral load in LRT samples at day 29**, n/N (%) | 1/9 (11.1%) | 0/2 (0.0%) | 0/3 (0.0%) | 0/2 (0.0%) | 1/2 (50.0%) |

#### The DisCoVeRy Study Group:

##### The French DisCoVeRy Trial Management Team:

F Ader, Y Yazdanpanah, F Mentre, N Peiffer-Smadja, FX Lescure, J Poissy, L Bouadma, JF Timsit, B Lina, F Morfin-Sherpa, M Bouscambert, A Gaymard, G Peytavin, L Abel, J Guedj, C Andrejak, C Burdet, C Laouenan, D Belhadi, A Dupont, T Alfaiate, B Basli, A Chair, S Laribi, J Level, M Schneider, MC Tellier, A Dechanet, D Costagliola, B Terrier, M Ohana, S Couffin-Cadiergues, H Esperou, C Delmas, J Saillard, C Fougerou, L Moinot, L Wittkop, C Cagnot, S Le Mestre, D Lebrasseur-Longuet, V Petrov-Sanchez, A Diallo, N Mercier, V Icard, B Leveau, S Tubiana, B Hamze, A Gelley, M Noret, E D'Ortenzio, O Puechal, C Semaille.

**The DisCoVeRy Steering Committee (members who are not listed in other groups):**

T Welte, JA Paiva, M Halanova, MP Kieny

**ANRS (France Recherche Nord&Sud SIDA-HIV Hépatites), Paris, France:** E Balssa, C Birkle, S Gibowski, E Landry, A Le Goff, L Moachon, C Moins, L Wadouachi, C Paul

**Centre Hospitalier Annecy Genevois, France:** D Bougon

**Centre Hospitalier de Cayenne Andrée Rosemon, Cayenne, France:** F Djossou, L Epelboin

**Centre Hospitalier Universitaire de Nice, France:** J Dellamonica, CH Marquette

**Centre Hospitalier Régional de Metz-Thionville, France:** C Robert

**Centre Hospitalier Régional Universitaire de Nancy, France:** S Gibot

**Centre Hospitalier de Tourcoing, France:** E Senneville, V Jean-Michel

**Centre Hospitalier Universitaire de Amiens, France:** Y Zerbib

**Centre Hospitalier Universitaire de Besançon, France:** C Chirouze

**Centre Hospitalier Universitaire de Bordeaux, France:** A Boyer, C Cazanave, D Gruzon, D Malvy

**Centre Hospitalier Universitaire de Dijon, France:** P Andreu, JP Quenot

**Centre Hospitalier Universitaire de Grenoble Alpes, France:** N Terzi

**Centre Hospitalier Universitaire de Lille, France:** K Faure

**Centre Hospitalier Universitaire de Martinique, Fort-de-France, France:** C Chabartier

**Centre Hospitalier Universitaire de Montpellier, France:** V Le Moing, K Klouche

**Centre Hospitalier Universitaire de Lyon, France:** T Ferry, F, Valour

**Centre Hospitalier Universitaire de Nantes, France:** B Gaborit, E Canet, P Le Turnier, D Boutoille

**Centre Hospitalier Universitaire de Reims, France:** F Bani-Sadr

**Centre Hospitalier Universitaire de Rennes, France:** F Benezit, M Revest, C Cameli, A Caro, MJ Ngo Um Tegue, Y Le Tulzo, B Laviolle, F Laine

**Centre Hospitalier Universitaire de Saint-Étienne, France:** G Thiery

**Centre Hospitalier Universitaire de Strasbourg, France:** F Meziani, Y Hansmann, W Oulehri, C Tacquard

**Centre Hospitalier Universitaire de Toulouse, France:** F Vardon-Bounes, B Riu-Poulenc, M Murris-Espin

**Centre Hospitalier Universitaire de Tours, France:** L Bernard, D Garot

**Groupe Hospitalier de Mulhouse Sud Alsace, France:** O Hinschberger

**Hospices Civils de Colmar, France:** M Martinot

**Groupe Hospitalier de Paris Saint Joseph, Paris, France:** C Bruel, B Pilmis

**Hôpital Avicenne, Assistance Publique – Hôpitaux de Paris, France:** O Bouchaud

**Centre Hospitalier Universitaire de Nîmes, France:** P Loubet, C Roger

**Hôpital Bicêtre, Assistance Publique – Hôpitaux de Paris, France:** X Monnet, S Figueiredo

**Hôpital Bichat - Claude Bernard, Assistance Publique – Hôpitaux de Paris, France:** V Godard

**Hôpital Cochin, Assistance Publique – Hôpitaux de Paris, France:** JP Mira, M Lachatre, S Kerneis

**Hôpital Delafontaine, Saint-Denis, France:** J Aboab, N Sayre, F Crockett

**Hôpital Européen Georges-Pompidou, Assistance Publique – Hôpitaux de Paris, France:** D Lebeaux, A Buffet, JL Diehl, A Fayol, JS Hulot, M Livrozet

**Hôpital Henri-Mondor, Assistance Publique – Hôpitaux de Paris, France:** A Mekontso-Dessap

**Hôpital d'Instruction des Armées Bégin, Saint Mandé, France:** C Ficko

**Hôpital Marie Lannelongue, Le Plessis Robinson, France:** F Stefan, J Le Pavec

**Hôpital de la Pitié-Salpêtrière, Assistance Publique – Hôpitaux de Paris, France:** J Mayaux

**Hôpital Saint-Antoine, Assistance Publique – Hôpitaux de Paris, France:** H Ait-Oufella

**Hôpital Saint-Louis, Assistance Publique – Hôpitaux de Paris, France:** JM Molina

**Hôpital Tenon, Assistance Publique – Hôpitaux de Paris, France:** G Pialoux, M Fartoukh

**Hospices Civils de Lyon, France:** J Textoris

**Inserm U1018, Université Paris Saclay, CESP, Paris, France:** M Brossard, A Essat

**Inserm US19-Sc10, Université Paris Saclay, Villejuif, France:** E Netzer, Y Riault, M Ghislain

**Sorbonne Université, INSERM U1136, Institut Pierre Louis d'Épidémiologie et de Santé Publique (IPLESP), Paris, France:** L Beniguel, M Genin

**Cliniques Universitaires de Saint Luc, Bruxelles, Belgique:** L Belkhir

**Centre Hospitalier Régional de la Citadelle, Liège, Belgique:** A Altdorfer, V Fraipont

**Centro Hospital Universitário de Lisboa Norte, Hospital de Santa Maria, Portugal:** S Braz, JM Ferreira Ribeiro

**Centro Hospitalar Universitário São João de Porto, Portugal:** JA Paiva, R Roncon Alburqueque

**Hôpitaux Robert Schuman, Luxembourg:** M Berna

**Luxembourg Institute of Health, Strassen, Luxembourg:** M Alexandre

**Kepler Universitätsklinikum Linz, Linz, Austria:** B Lamprecht

**Paracelsus Medical University Salzburg, SCRI-CCCIT and AGMT, Austria:** A Egle, R Greil

**AGMT Arbeitsgemeinschaft Medikamentöse Tumortherapie, Salzburg, Austria:** R Greil

**Medizinische Universität Innsbruck, Innsbruck, Austria:** M Joannidis

#### Acknowledgements

We gratefully acknowledge the members of the Data Safety Monitoring Board: Stefano VELLA (chair), Guanhua DU, Donata MEDAGLINI, Mike JACOBS, Sylvie VAN DER WERF, Stuart

POCOCK, Phaik Yeong CHEAH, Karen BARNES, Patrick YENI, and the independent statistician Tim COLLIER.

We gratefully acknowledge the chief executive officer of Inserm: Dr. Gilles BLOCH.

We acknowledge the RENARCI network: Marion NORET, Pierre TATTEVIN and Albert SOTTO.

We acknowledge the outstanding support of the Clinical Investigation Centers: Pierre-Olivier GIRODET (CIC1401, Inserm, CHU Bordeaux), Dominique DEPLANQUE (CIC1403, Inserm, CHRU LIlle), Jean-Luc CRACOWSKI (CIC1406, Inserm, CHU Grenoble), Michel OVIZE (CIC1407, Inserm, Hospices Civil de Lyon), Bernard TARDY (CIC1408, Inserm, CHU Saint-Etinne), Eric RENARD (CIC1411, Inserm, CHU Montpellier), Jean-Noël TROCHU (CIC1413, Inserm, CHU Nantes), Bruno LAVIOLLE (CIC1414, Inserm, CHU Rennes), Wissam EL HAGE (CIC1415, Inserm, CHU Tours), Odile LAUNAY (CIC1417, Inserm, APHP Cochin), Jean-Sébastien HULOT (CIC1418, Inserm, APHP HEGP), Jean-Christophe CORVOL (CIC1422, Inserm, APHP Pitié-Salpétriêre), Mathieu NACHER (CIC1424, Inserm, CH André Rosemon), André CABIE (CIC1424, Inserm, CHU de la Martinique), Xavier Duval (CIC1425, Inserm, APHP Bichat), Philippe LE CORVOISIER Philippe (CIC1430, Inserm, APHP Mondor), Emmanuel HAFFEN (CIC1431, Inserm, CHU Besançon), Marc BARDOU (CIC1432, Inserm, CHU Dijon), Patrick ROSSIGNOL (CIC1433, Inserm, CHU Nancy), Catherine SCHMIDT-MUTTER (CIC1434, Inserm, CHRU Strasbourg), Olivier RASCOL (CIC1436, Inserm, CHU Toulouse).

We acknowledge the pharmacists: Laure LALANDE (Hospices Civils de Lyon), Marine AUSSEDAT (Hospices Civils de Lyon), Jennifer LE GRAND (APHP Bichat, PARIS), Laura KRAMER (APHP Bichat, PARIS), Sylvie BRICE (CHRU Lille), Sayah MEGUIG (CHRU Lille), Laurent FLET (CHU Nantes), Martine TCHING-SIN (CHU Nantes), Audrey LEHMANN (CHU Grenoble), Sophie Cerana (CHU Grenoble), Mélanie Minovès

(CHU Grenoble), Gwenaël MONNIER (CHU Saint Etienne), Guillaume BECKER (CHRU Strasbourg), Anne HUTT-CLAUSS (CHRU Strasbourg), Sylvia WEHRLEN-PUGLIESE (CHU Nice), Carine GHIONDA (CHU Nice), Marie-Christine RIGAULT (CHU Nice), Christelle BOCZEK (CHU Nice), Justine BELLEGARDE (CHU Nice), Audrey CASTET-NICOLAS (CHU Montpellier), Jean GALLOT-LAVALLEE (CHU Montpellier), Sophie JUAREZ (CHU Montpellier), Valérie CARRE (CHU Montpellier), Fanny CHARBONNIER-BEAUPEL(APHP Pitié-Salpêtrière, Carole METZ (APHP Pitié-Salpêtrière), MURIEL CARVALHO VERLINDE (APHP Mondor), ALAKI THIEMELE (APHP Mondor), delphine LE FEBVRE DE NAILLY (APHP Mondor), Magali FARINES RAFFOUL (CH Annecy Genevois), Franck GUERIN (CH Annecy Genevois), Bénédicte BONNARD (CH Annecy Genevois), Corinne PERNOT (CHU Dijon), Amélie CRANSAC (CHU Dijon), Julie JAMBON (CHU Dijon), Julien MORLET (CHU Dijon), Clotilde Le Tiec (APHP Bicêtre), Aurélie Barrail Tran (APHP Bicêtre), Renée Flore ROY EMA (APHP Bicêtre), Grégory Rondelot (CHR Metz Thionville), Julie Bernez (CHR Metz Thionville), Alexia Hermitte (CHR Metz Thionville), Hélène Perraud (CHR Metz Thionville), Julien Voyat (CHR Metz Thionville), Laurence Ferrier (CHR Metz Thionville), Julie Alips (CHR Metz Thionville), Sarah Di-Filippo (CHR Metz Thionville), Florence Beringuer (CHR Metz Thionville), Michel PREVOT (CHRU Nancy), Sophie MORICE (CHRU Nancy), Brigitte SABATIER (APHP HEGP), Dominique ROUSSEAU (APHP HEGP), Marie KROEMER (CHRU Besançon), Julie VARDANEGA (CHRU Besançon), Magalie VIEILLE (CHRU Besançon), Karen GEORGE (GHR Mulhouse Sud Alsace), Marion TISSOT (GHR Mulhouse Sud Alsace), Laetitia ARNOUX (GHR Mulhouse Sud Alsace), Marjorie BRIMBOEUF (GHR Mulhouse Sud Alsace), Philippe BENOIT (CHU Reims), Frédéric EYVRARD (CHU Toulouse), Caroline SORLI (CHU Toulouse), Corinne GUERIN (APHP Cochin), Jeremie ZERBIT (APHP Cochin), MERIAM JARDIN SUZCS (HPJS), SOPHIE RENET (HPJS), Alix DE CHEVIGNY (HPJS),

Anne Daguenel-Nguyen (APHP Saint-Antoine), Clementine Mayala Kanda (APHP Saint-Antoine), Catherine HAMON-BOUER (CHU Rennes), Pascal LE CORRE (CHU Rennes), Valentine JASPART-LE DU (CHU Rennes), Roza WILSON (CHU Rennes), Sophie BODDAERT (CHU Amiens), Florian SMAGGHE (CHU Amiens), Mickael DELAUNAY (CHU Amiens), Véronique Dubar (CH Tourcoing), Adeline Danielou (CH Tourcoing), Ghezzoul BELLABES (CHU Bordeaux), Salima ALLAF (APHP Tenon), Jean-Baptiste PAIN (APHP Tenon), Nina DURET-AUPY (CHU Tours), Hélène BOURGOIN (CHU Tours), Thierno DIEYE (Hopital Delafontaine), Anais RAZUREL (Hopital Delafontaine), Jean-Louis LAMAIGNERE (CHU Martinique), Aurélie RISAL (CHU Martinique), Amandine SGARIOTO (Hôpital des Armées Begin), Claire DAVOINE (APHP Hôpital Saint-Louis), Vanessa RATHOUIN (Hôpital Avicenne), Blaise NICAISE (CH Andrée Rosemon), Gregory Gaudillot (CHL, Luxembourg), Emmanuelle Andlauer-Vialette (CHL, Luxembourg), Anne OTTO (HRS, Luxembourg), Francoise FRANTZEN (HRS, Luxembourg), Nathalie BART (HRS, Luxembourg), Martin Wolkersdorfer (Landeskrankenhaus, Salzburg), Astrid Vandorpe (Hôpital Erasme, Bruxelles), Axelle Scarnière (Hôpital Erasme, Bruxelles), Kristel Marquez (Hôpital Erasme, Bruxelles), Nathalie Mathonet (Hôpital Erasme, Bruxelles), Robert Rugwiro (Hôpital Erasme, Bruxelles), Jarode Ducrot (Hôpital Erasme, Bruxelles), Florence Vandenberghe (Hôpital Erasme, Bruxelles), Alexa Jonas (CHR Citadelle, Liège), Valentine Lesenfant (CHR Citadelle, Liège), Blaise Delhauteur (CHR Citadelle, Liège), Annick Peters (CHR Citadelle, Liège), Thi Nguyen (CHR Citadelle, Liège), Jean-François Gouders (CHR Citadelle, Liège), Grégory Opeind (CHR Citadelle, Liège), Adeline GAILLET (Cliniques universitaires Saint Luc, Bruxelles), Ana Carrondo (CHU Lisboa Norte, Hospital Santa Maria, Lisboa), Daniela Ramos (CHU Lisboa Norte, Hospital Santa Maria, Lisboa), Vanessa Côdea (CHU Lisboa Norte, Hospital Santa Maria, Lisboa), Raquel Carneiro CHU Lisboa Norte,

Hospital Santa Maria, Lisboa), Ana Lima (CHU Lisboa Norte, Hospital Santa Maria, Lisboa), Tatiana Bento (CHU Lisboa Norte, Hospital Santa Maria, Lisboa).

We acknowledge the following individuals from the French Data Management Team of Discovery:

Drug supply: Johanna GUILLON, Anne-Marie TABURET.

Inserm-Transfert: Pascale AUGÉ, Mireille CARALP, Nathalie DUGAS, Florence CHUNG, Julia LUMBROSO.

Clinical Research Unit CHU Bichat-Claude Bernard: Camille COUFFIGNAL.

We thank all the staff members involved in data monitoring.

We thank the COVID-19 Partners Platform and Hervé Le Nagard for data hosting.

We thank Theradis PHARMA, Cagnes-sur-Mer, France.

This study was conducted with the support of GILEAD, SANOFI, MERCK and ABBVIE that provided the study drugs.

### CONSORT 2010 checklist of information to include when reporting a randomised trial*

| Section/Topic | Item No | Checklist item | Reported on page No |
|---|---|---|---|
| **Title and abstract** | | | |
| | 1a | Identification as a randomised trial in the title | 1 |
| | 1b | Structured summary of trial design, methods, results, and conclusions (for specific guidance see CONSORT for abstracts) | 8 |
| **Introduction** | | | |
| Background and objectives | 2a | Scientific background and explanation of rationale | 10 |
| | 2b | Specific objectives or hypotheses | 10 |
| **Methods** | | | |
| Trial design | 3a | Description of trial design (such as parallel, factorial) including allocation ratio | 11, 12 |
| | 3b | Important changes to methods after trial commencement (such as eligibility criteria), with reasons | NA |
| Participants | 4a | Eligibility criteria for participants | 11 |
| | 4b | Settings and locations where the data were collected | 15 |
| Interventions | 5 | The interventions for each group with sufficient details to allow replication, including how and when they were actually administered | 12 |
| Outcomes | 6a | Completely defined pre-specified primary and secondary outcome measures, including how and when they were assessed | 12, 13 |
| | 6b | Any changes to trial outcomes after the trial commenced, with reasons | 13 |
| Sample size | 7a | How sample size was determined | 13 |
| | 7b | When applicable, explanation of any interim analyses and stopping guidelines | 13 |
| Randomisation: | | | |
| Sequence generation | 8a | Method used to generate the random allocation sequence | 13 |
| | 8b | Type of randomisation; details of any restriction (such as blocking and block size) | 13 |
| Allocation concealment mechanism | 9 | Mechanism used to implement the random allocation sequence (such as sequentially numbered containers), describing any steps taken to conceal the sequence until interventions were assigned | 13 |
| Implementation | 10 | Who generated the random allocation sequence, who enrolled participants, and who assigned participants to interventions | 13 |
| Blinding | 11a | If done, who was blinded after assignment to interventions (for example, participants, care providers, those assessing outcomes) and how | 11 |
| | 11b | If relevant, description of the similarity of interventions | NA |

| | | | |
|---|---|---|---|
| Statistical methods | 12a | Statistical methods used to compare groups for primary and secondary outcomes | 14 |
| | 12b | Methods for additional analyses, such as subgroup analyses and adjusted analyses | 14 |
| **Results** | | | |
| Participant flow (a diagram is strongly recommended) | 13a | For each group, the numbers of participants who were randomly assigned, received intended treatment, and were analysed for the primary outcome | 15 |
| | 13b | For each group, losses and exclusions after randomisation, together with reasons | 15 |
| Recruitment | 14a | Dates defining the periods of recruitment and follow-up | 15 |
| | 14b | Why the trial ended or was stopped | 13 |
| Baseline data | 15 | A table showing baseline demographic and clinical characteristics for each group | 23 |
| Numbers analysed | 16 | For each group, number of participants (denominator) included in each analysis and whether the analysis was by original assigned groups | 23 |
| Outcomes and estimation | 17a | For each primary and secondary outcome, results for each group, and the estimated effect size and its precision (such as 95% confidence interval) | 23 |
| | 17b | For binary outcomes, presentation of both absolute and relative effect sizes is recommended | 23 |
| Ancillary analyses | 18 | Results of any other analyses performed, including subgroup analyses and adjusted analyses, distinguishing pre-specified from exploratory | 16, 17, 28 |
| Harms | 19 | All important harms or unintended effects in each group (for specific guidance see CONSORT for harms) | 16, 17, 28 |
| **Discussion** | | | |
| Limitations | 20 | Trial limitations, addressing sources of potential bias, imprecision, and, if relevant, multiplicity of analyses | 19 |
| Generalisability | 21 | Generalisability (external validity, applicability) of the trial findings | 18, 19 |
| Interpretation | 22 | Interpretation consistent with results, balancing benefits and harms, and considering other relevant evidence | 17, 18 |
| **Other information** | | | |
| Registration | 23 | Registration number and name of trial registry | 11 |
| Protocol | 24 | Where the full trial protocol can be accessed, if available | |
| Funding | 25 | Sources of funding and other support (such as supply of drugs), role of funders | |

*We strongly recommend reading this statement in conjunction with the CONSORT 2010 Explanation and Elaboration for important clarifications on all the items. If relevant, we also recommend reading CONSORT extensions for cluster randomised trials, non-inferiority and equivalence trials, non-pharmacological treatments, herbal interventions, and pragmatic trials. Additional extensions are forthcoming: for those and for up to date references relevant to this checklist, see www.consort-statement.org.
